# Relationship between nursing home COVID-19 outbreaks and staff neighborhood characteristics

**DOI:** 10.1101/2020.09.10.20192203

**Authors:** Karen Shen

## Abstract

The COVID-19 pandemic has been particularly deadly for residents of nursing homes and other long-term care facilities. This paper analyzes COVID-19 deaths at nursing homes during the first wave of the pandemic in the United States during the spring and early summer 2020. By combining data on facility-level COVID-19 deaths during this period with data on the neighborhoods where nursing home staff reside for a sample of eighteen states, this paper finds that staff neighborhood characteristics were a large and significant predictor of COVID-19 nursing home deaths. Even after controlling for the county where a facility is located, one standard deviation increases in average staff neighborhood (Census tract) population density, public transportation use, and non-white share were associated with 1.3 (p < .001), 1.4 (p<.001), and 0.9 (p<.001) additional deaths per 100 beds, respectively. These effects are larger than all facility management or quality variables, and larger than the effect of the nursing home’s own neighborhood characteristics. These results suggest COVID-19 outbreaks in staff communities can have large consequences for the facilities where they work, even in highly-rated facilities, and that disparities in nursing home outbreaks may be related to differences in the types of neighborhoods nursing home staff live in.

## Introduction

The COVID-19 pandemic’s overwhelming impact on nursing homes in the United States and worldwide has been well-documented [1]. As of November 22, 2020, there had been 256,597 deaths from COVID-19 nationwide and the Center for Medicare and Medicaid Services had recoded 72,642 deaths among nursing home residents. This number would already imply that 28% of deaths have been among nursing home residents; however, due to the fact that federal reporting was optional before May 10, it is likely to be a significant underestimate [2].

Numerous hypotheses have emerged about what factors affected the vulnerability of facilities to infection, with some pointing to poor management or infection control procedures, and others to specific actions and policies such as the timing of when nursing homes became locked down to visitors, state policies governing the transfer of recovering COVID-positive patients to nursing homes, and the supply of personal protective equipment (PPE) and testing [3][4]. Another possibility is that outbreaks are unpredictable and unpreventable and that luck and geography, rather than factors under a nursing home operator or policymaker’s control, largely determined which facilities saw outbreaks and which did not.

This study uses an eighteen-state sample of nursing homes to study facility-level determinants of COVID-19 deaths. The focus is on within-county differences in facility-level deaths (scaled by the number of beds in each facility) during the first wave of the pandemic in the spring and early summer of 2020. Using a novel approach to measure characteristics of the neighborhoods where each nursing home’s staff members live, this paper provides new evidence that the local geography of where nursing home staff live is a strong predictor of outbreak sizes, even within counties. Specifically, data from the Census Bureau on commuting patterns at the census-tract level is used to show that facilities whose staff live in denser and less white neighborhoods with higher public transportation use are significantly more likely to have larger outbreaks compared to other facilities in the same county. By contrast, and consistent with other earlier studies [5], this study finds no relationship between a facility’s star rating and the number of resident deaths from COVID-19 during the first wave of the pandemic.

These results suggest the possibility that the spread of COVID-19 in staff communities is an important transmission mechanism driving facility outbreaks, and that controlling community spread may be of first-order importance in attempts to protect nursing homes. To support this hypothesis, a subsample analysis is conducted on eight states that provided local infection rate data at the time of data collection. This analysis shows that nursing home outbreak sizes are highly correlated with the average COVID-19 case rate in staff towns (more so than of the nursing home’s town itself), and that controlling for the average staff town case rate is able to explain some of the effect of staff neighborhood characteristics.

This evidence implies that the exposure of a facility’s staff to the virus in their local communities, which can likely be proxied well by neighborhood population density, public transportation use, and non-white share, is a strong predictor of facility outbreak size. Our final analysis investigates what types of facilities are likely to have high staff neighborhood exposure within a county. While lower quality ratings and wages are somewhat correlated with high staff exposure, the strongest correlation is with the proportion of a nursing home’s residents who are non-white. These results thus offer a mechanism to explain the differential impact of the pandemic on facilities with more non-white residents that has been noted in the media and other studies [6, 7].

## Methods

### Data

The universe of study is all Medicare and Medicaid-certified nursing homes in the eighteen sample states. Data on the addresses, most recent star ratings, ownership, occupancy rate, and prior infection control-related violations are obtained from the Nursing Home Compare database. Other administrative data on chain ownership, resident demographics, and wages come from the OSCAR/CASPER data, the LTC Focus Database, and the HCRIS data, respectively.

Lists of nursing homes that have had COVID-19 cases were collected from eighteen state government websites between 7/5 and 7/10 and matched to the administrative data using fuzzy-matching and geocoding techniques. The eighteen states were selected primarily due to data availability, and having large numbers of deaths from COVID-19. State data is needed because the federal data did not require facility reporting before May 8, 2020, and is therefore missing a significant number of deaths. Specifics of this selection and differences across states in reporting practices are discussed in the supplementary material. Data on population case rates at fine geographies (town, zip or tract) was also collected from a subsample of eight states that released this data.

The Longitudinal Employer-Household Dynamics Origin-Destination Employment Statistics (LODES) data was used to measure which neighborhoods each facility’s staff live in. This data provides counts of employment for every work and home census block pair in three large industry groupings (“goods producing”, “trade/transportation/utilities”, “all other services”). Staff neighborhood measures were constructed using all workers on the nursing home’s census block in the “all other services’” category. Six percent of facilities with unrealistically low numbers of workers in this category were dropped. The data is a snapshot of all workers on April 1, 2017 and is thus unlikely to capture the exact neighborhoods of nursing home workers because of worker turnover or moves, but is likely to be representative of the type of neighborhoods in which the nursing home’s workers are likely to reside. Each worker’s home census tract is then matched to data from the American Community Survey (ACS) 5-year estimates (2014-2018) on tract-level population density, poverty, income, and use of public transportation, and employment by industry and occupation. The employment data is used to compute a predicted share of “frontline workers,” following other literature. [8, 9] Finally, weighted averages of these characteristics are taken at the facility-level for all tracts where staff live to obtain the final measures of facility staff exposures.

### Statistical Analyses

The main analyses are ordinary least squares regressions where the dependent variable is the cumulative number of reported COVID-19 deaths at a facility as of early July per 100 beds. Deaths are used because they are likely to be less dependent on testing and more consistently measured across facilities. Independent continuous variables are scaled to have a standard deviation of 1 to allow for comparisons of effect sizes, while binary variables are unchanged. regressions contain county fixed effects. In general, the independent variables chosen are standard to the literature, and cross-correlations of all independent variables of interest were computed in order to avoid issues of multi-collinearity. Regression analyses were conducted using Stata 16.1. The facility-level data on deaths and tract-level neighborhood characteristics used in this paper are available at DOI 10.5281/zenodo.4308760.

## Results

Figure 1 shows total deaths and deaths among nursing home residents from COVID-19 per week in the US from the beginning of the pandemic through the end of November. Weekly nursing home deaths are plotted using the CMS data, which is available beginning the week ending May 31. The graph shows that for the period where the federal data is available, nursing homes have accounted for a large share of COVID-19 deaths, particularly near the beginning of the sample (the end of the first wave). These trends emphasize both the importance of studying the first wave of the pandemic, which has thus far been the deadliest period overall and particularly for nursing homes, and the need to use an alternative source of nursing home deaths to the CMS data to do so.

**Figure 1.**
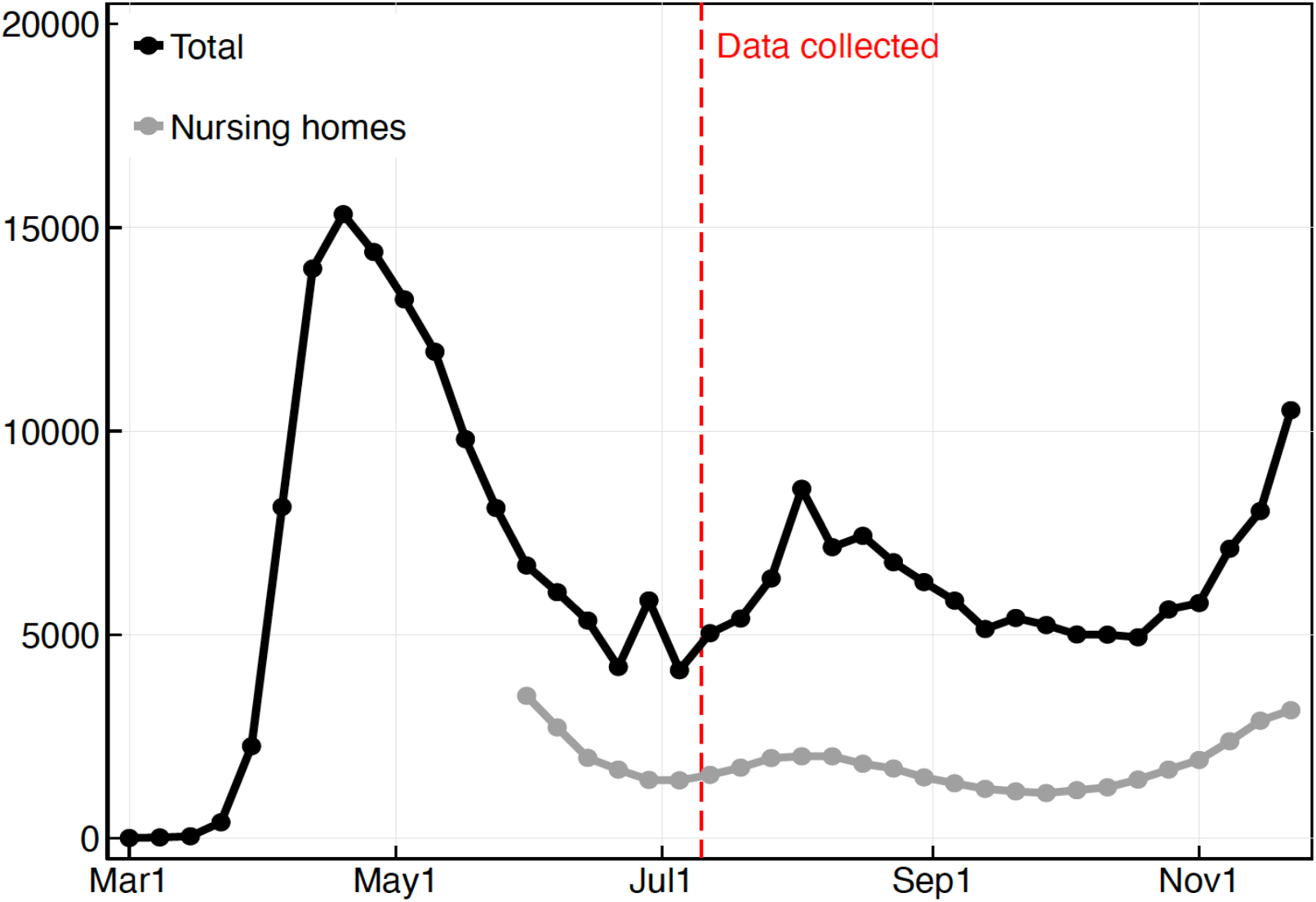
Weekly deaths from COVID-19 in the United States, total and in nursing homes. The black line shows weekly deaths from COVID-19 from the New York Times database, and the grey line shows weekly deaths in nursing homes using the CMS database.

Figure 2 introduces the eighteen state sample used in this paper. The sample includes states from each region of the country. The impact of the first wave of the pandemic on these states varied substantially. Figure 2 summarizes the main outcome variable (facility deaths per 100 beds) by state. Overall, at the time of the data, the average nursing home in this sample had experienced 3.7 deaths per bed. For some states in the Northeast (MA, NJ, CT, RI), this number was more than 8 deaths per bed, while some states in other regions (WV, FL, NC, SC, NV) had experienced fewer than 2 deaths per bed. This variation is roughly in line with variation in total deaths during the first wave of the pandemic, but some of these differences may be because states varied in their reporting requirements. Since the main analysis controls for each facility’s county, these differences in reporting should have less impact on the main results.

**Figure 2.**
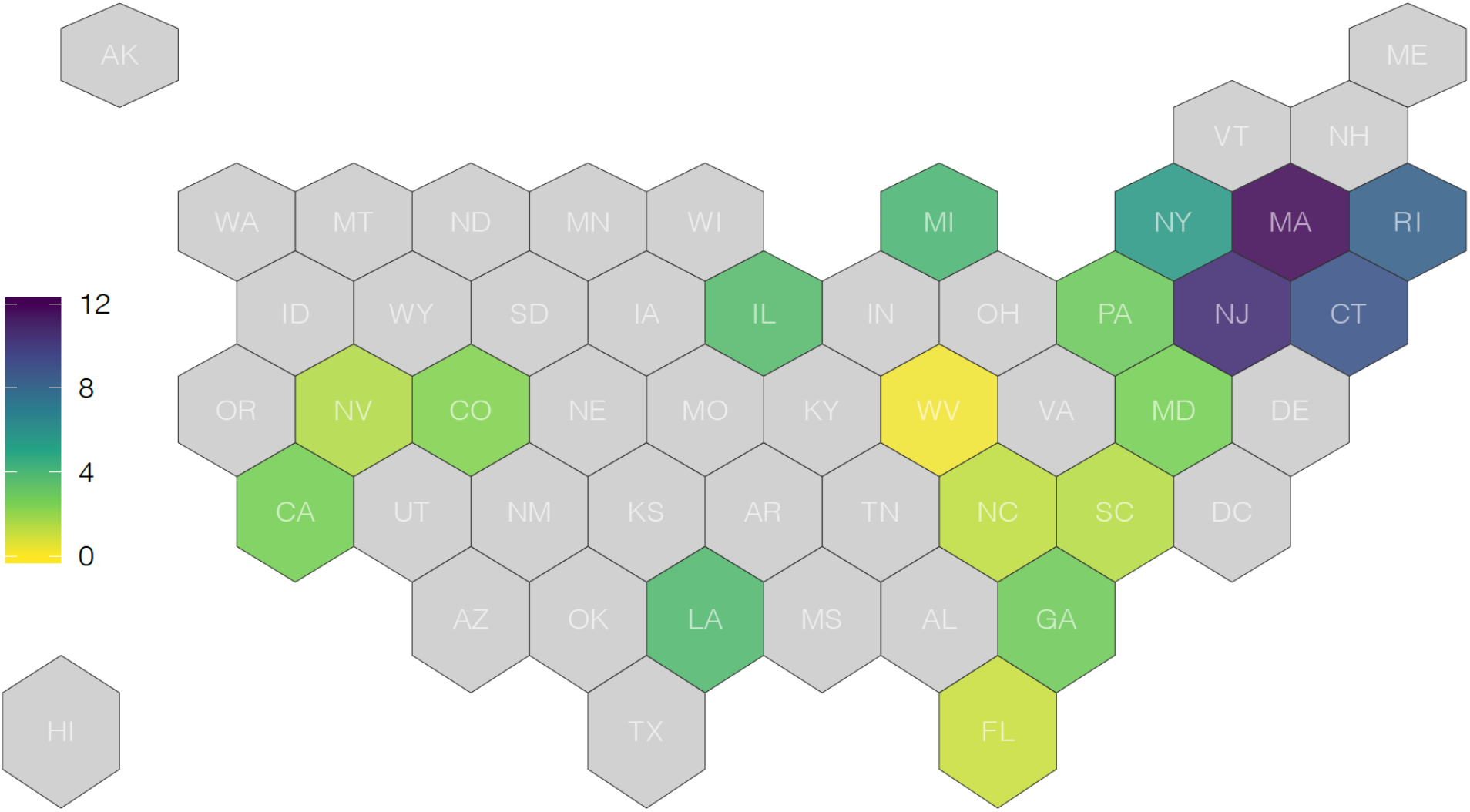
Average facility deaths per bed in sample states as of early July 2020. States are shaded according to the average number of deaths per bed at nursing homes in each state. States not included sample are in light grey. N=6,132.

Figure 3 shows how the number of deaths per bed at a facility is related to a set of variables of interest. Nursing home outbreak sizes are strongly related to the county infection rate (Panel A), as well as the number of beds in the facility (Panel B). Panel C plots average death rates by star rating and finds virtually no relationship between the two variables. The bottom row of Figure 3 shows the relationship of facility deaths to staff neighborhood characteristics. Nursing homes are grouped by the population density, public transportation use (share of workers who commute to work on public transportation), and non-white share of the census tract where their staff live (in blue) and the tract where they are located (in orange). Facility outbreaks are strongly associated with all three of these characteristics, with steeper slopes for staff neighborhoods than the nursing home neighborhood in each case.

**Figure 3.**
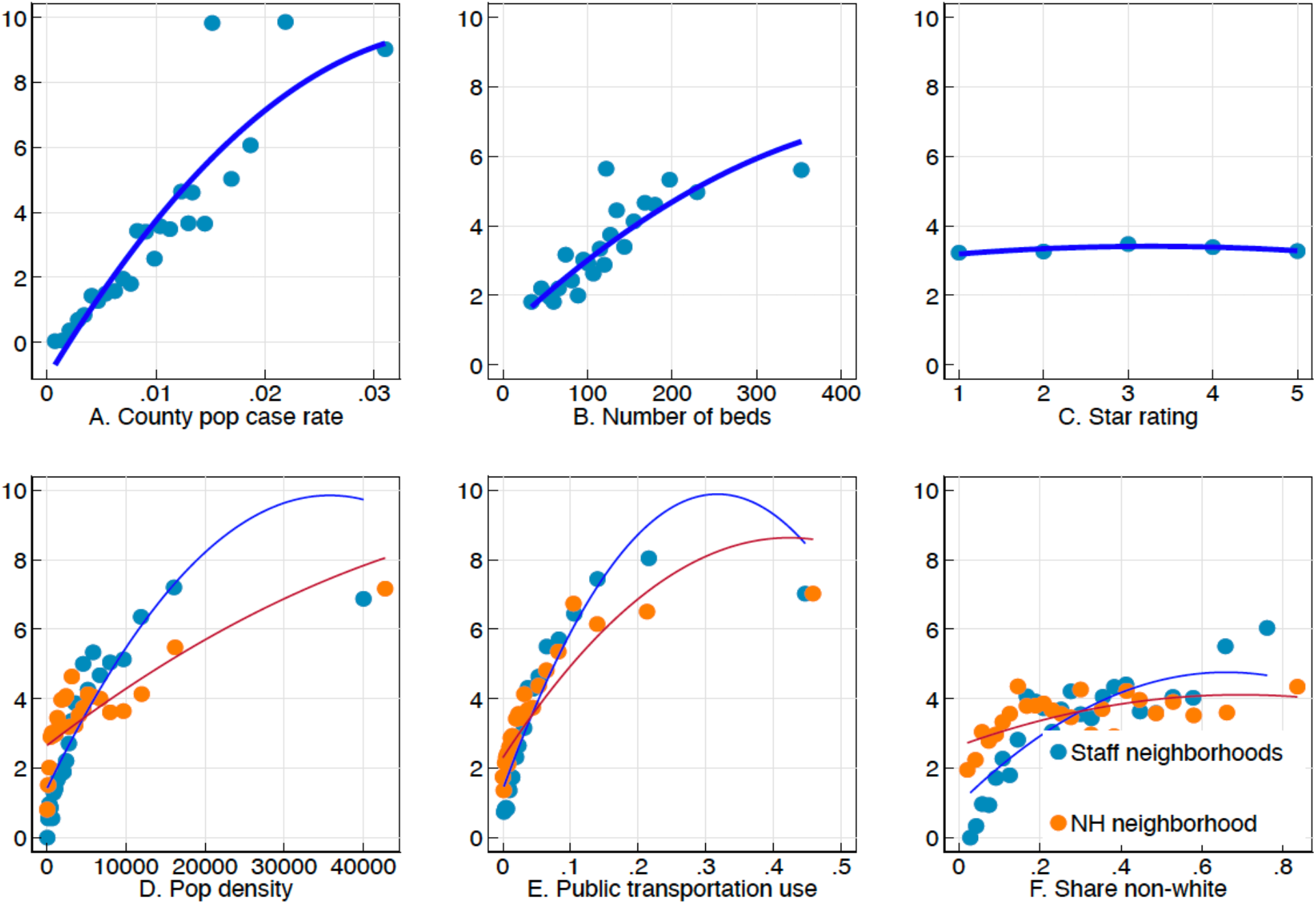
Relationship of facility deaths per bed with facility geography measures, size, and star rating. Each panel of the Figure shows average nursing home deaths per 100 beds on the y-axis binned by a different variable along the x-axis. Bins are of equal sizes, and the line represents a quadratic fit.

Table 1 presents models that jointly estimate the effect of facility characteristics and staff and nursing home neighborhood characteristics on facility deaths per bed. As of early July, there had been an average of 3.7 deaths per 100 beds across all facilities in the sample summary statistics for the independent variables are provided in S1 Table). The model in column (1) does not include any neighborhood characteristics, and shows that for-profit nursing homes are associated with an additional .53 deaths (SE .52, p=.007) and nursing homes that belong to chains are associated with an additional .35 deaths (SE .16, p=.033) per 100 beds. As seen above, even after scaling deaths by the number of beds in a facility, facility size continues to be an important factor in explaining death rates, as are occupancy rates, which may reflect both the mechanical effect of more residents per bed in the facility and potential crowding effects. On the other hand, there is no significant relationship of facility outbreak size with the star rating, prior infection control violations, the Medicaid share, or the non-white share after accounting for the other controls and county fixed effects.

**Table 1:** Effect of staff tract measures and own tract measures on facility deaths per bed.

|  | (1) | (2) | (3) | (4) | (5) | (6) |
| --- | --- | --- | --- | --- | --- | --- |
| Staff tract pop density |  | 1.307***<br>(0.331) |  |  |  |  |
| NH tract pop density |  | -0.139<br>(0.141) |  |  |  |  |
| Staff tract pubtrans use |  |  | 1.362***<br>(0.347) |  |  |  |
| NH tract pubtrans use |  |  | -0.0890<br>(0.177) |  |  |  |
| Staff tract share nonwhite |  |  |  | 0.857***<br>(0.235) |  |  |
| NH tract share nonwhite |  |  |  | -0.0589<br>(0.127) |  |  |
| Staff tract pov rate |  |  |  |  | 0.519**<br>(0.168) |  |
| NH tract pov share |  |  |  |  | -0.153<br>(0.0945) |  |
| Staff tract share frontline |  |  |  |  |  | 0.194<br>(0.201) |
| NH tract share frontline |  |  |  |  |  | -0.177<br>(0.111) |
| For-profit | 0.525**<br>(0.193) | 0.504**<br>(0.193) | 0.522**<br>(0.193) | 0.522**<br>(0.194) | 0.510**<br>(0.193) | 0.516**<br>(0.194) |
| Chain | 0.348*<br>(0.163) | 0.380*<br>(0.163) | 0.370*<br>(0.163) | 0.360*<br>(0.163) | 0.365*<br>(0.163) | 0.347*<br>(0.163) |
| Overall Rating | 0.0468<br>(0.0916) | 0.0474<br>(0.0915) | 0.0478<br>(0.0915) | 0.0529<br>(0.0917) | 0.0530<br>(0.0916) | 0.0471<br>(0.0916) |
| No prior infection viol. | 0.257<br>(0.196) | 0.260<br>(0.196) | 0.256<br>(0.196) | 0.249<br>(0.196) | 0.251<br>(0.196) | 0.260<br>(0.196) |
| Medicaid share | -0.0147<br>(0.0909) | 0.00419<br>(0.0909) | 0.000398<br>(0.0908) | -0.00875<br>(0.0908) | -0.0257<br>(0.0910) | -0.00771<br>(0.0915) |
| Resident share nonwhite | -0.0680<br>(0.118) | -0.171<br>(0.121) | -0.218<br>(0.124) | -0.282*<br>(0.138) | -0.161<br>(0.125) | -0.0574<br>(0.120) |
| Avg severity | -0.0811<br>(0.0852) | -0.0693<br>(0.0852) | -0.0548<br>(0.0853) | -0.0463<br>(0.0856) | -0.0517<br>(0.0858) | -0.0761<br>(0.0855) |
| Occupancy Rate | 0.681***<br>(0.0886) | 0.668***<br>(0.0885) | 0.673***<br>(0.0884) | 0.681***<br>(0.0885) | 0.676***<br>(0.0886) | 0.681***<br>(0.0886) |
| 25-50 beds | 0<br>(.) | 0<br>(.) | 0<br>(.) | 0<br>(.) | 0<br>(.) | 0<br>(.) |
| 50-100 beds | 0.486<br>(0.329) | 0.506<br>(0.328) | 0.538<br>(0.328) | 0.515<br>(0.328) | 0.498<br>(0.329) | 0.485<br>(0.329) |
| 100-150 beds | 1.047**<br>(0.334) | 1.066**<br>(0.333) | 1.107***<br>(0.333) | 1.093**<br>(0.334) | 1.062**<br>(0.333) | 1.048**<br>(0.334) |
| 150-200 beds | 1.688***<br>(0.369) | 1.670***<br>(0.369) | 1.712***<br>(0.369) | 1.679***<br>(0.369) | 1.675***<br>(0.369) | 1.676***<br>(0.369) |
| 200+ beds | 1.156**<br>(0.410) | 1.114**<br>(0.410) | 1.164**<br>(0.409) | 1.137**<br>(0.410) | 1.151**<br>(0.410) | 1.152**<br>(0.410) |
| Constant | 2.164***<br>(0.344) | 2.181***<br>(0.344) | 2.132***<br>(0.344) | 2.166***<br>(0.344) | 2.164***<br>(0.344) | 2.172***<br>(0.344) |
| Fixed Effects | County | County | County | County | County | County |
| Depvar mean | 3.735 | 3.735 | 3.735 | 3.735 | 3.735 | 3.735 |
| Adj R2 | 0.29 | 0.29 | 0.29 | 0.29 | 0.29 | 0.29 |
| N | 6132 | 6132 | 6132 | 6132 | 6132 | 6132 |

This table reports OLS regression estimates of facility deaths per 100 beds on a collection of facility characteristics and county fixed effect. All continuous variables are normalized to have a standard deviation of 1. Standard errors are reported in parentheses. Significance: \* $p < .05$ , \*\* $p < .01$ , \*\*\* $p < .001$

Columns (2)-(6) add characteristics of the staff and nursing home neighborhoods. The population density (column 2), public transportation use (column 3), and non-white share (column 4) of staff neighborhoods are all highly and statistically significantly associated with facility deaths per bed. For example, a one standard deviation increase in the average staff neighborhood population density is associated with an additional 1.3 deaths per 100 beds (p<.001, column 2); and comparably sized changes in staff neighborhood public transportation use, nonwhite share, and poverty rate are associated with an additional 1.4 (p<.001), 0.9 (p<.001), and 0.5 (p<.001) deaths per 100 beds, respectively (columns 3-5). In all of these cases, the same characteristic measured for the nursing home’s census tract is not statistically different from zero when the staff neighborhood variable is included. Column 6 shows no effect of the share of workers who are predicted to be frontline workers. The magnitudes of the effects in columns (2)-(4) are larger than any of the effects of other facility characteristics except facility size, suggesting that local staff geography may be an incredibly important factor in determining facility outbreaks. Most of the relationships from column (1) are unchanged by the inclusion of these neighborhood variables: size, for-profit status, and chain status continue to have significant effects on facility outbreaks. The one exception is the resident non-white share, which becomes more negatively correlated with facility deaths once staff neighborhood characteristics are added in columns (2)-(5).

A likely interpretation of the results in Table 1 is that nursing home staff members are an important source of infection, and that the identified neighborhood characteristics are good proxies for the level of community spread in a neighborhood. Unfortunately, few states report infection rates at the neighborhood level (counties are the lowest level of geography that is available nationally). Table 2 uses a subsample of eight states that provide more local data on case rates to test this interpretation. Columns (1) and (3) reproduce columns (3) and (4) (public transportation use and non-white share) from Table 1 for this smaller subsample, since these variables exhibited the strongest associations with facility infection in the full sample (because population density is highly correlated with public transportation use (ρ=.92), it is omitted for the sake of brevity). Both effects are still positive and highly significant predictors of facility death rates in this smaller subsample. Columns (2) and (4) add measures of the population case rate of COVID-19 in staff towns and the nursing home’s town (again normalized to have standard deviations of 1). A one standard deviation in the average infection rate of staff towns is associated with an additional 2.2 (p<.001, column 2) or 2.4 deaths per 100 beds (p<.001, column 4) at a facility. The infection rate of the nursing home’s town is associated with a smaller, but still large, increase in deaths. After including these measures, the estimated effects on staff neighborhood public transportation use and staff neighborhood non-white share are significantly reduced, suggesting that it is quite possible that those effects operate through differences in community-level infection.

**Table 2:** Relationship of nursing home deaths with local infection rates in staff and nursing home neighborhoods.

|  | (1) | (2) | (3) | (4) |
| --- | --- | --- | --- | --- |
| Staff tract pubtrans use | 1.412***<br>(0.303) | 0.924**<br>(0.316) |  |  |
| Staff tract share nonwhite |  |  | 1.345***<br>(0.354) | 0.604<br>(0.380) |
| Staff town avg case rate |  | 2.190***<br>(0.452) |  | 2.330***<br>(0.466) |
| NH town case rate |  | 0.546***<br>(0.159) |  | 0.532***<br>(0.160) |
| For-profit | 0.145<br>(0.361) | 0.277<br>(0.357) | 0.208<br>(0.362) | 0.315<br>(0.358) |
| Chain | 0.510<br>(0.302) | 0.480<br>(0.298) | 0.422<br>(0.302) | 0.420<br>(0.298) |
| Overall Rating | -0.0483<br>(0.173) | -0.0525<br>(0.171) | -0.0165<br>(0.174) | -0.0372<br>(0.171) |
| No prior infection viol. | 0.367<br>(0.355) | 0.404<br>(0.352) | 0.369<br>(0.356) | 0.416<br>(0.352) |
| Medicaid share | 0.0245<br>(0.183) | 0.0126<br>(0.181) | 0.00831<br>(0.183) | -0.000745<br>(0.181) |
| Resident share nonwhite | -0.230<br>(0.220) | -0.481*<br>(0.222) | -0.383<br>(0.243) | -0.507*<br>(0.241) |
| Avg severity | 0.369*<br>(0.150) | 0.402**<br>(0.149) | 0.387*<br>(0.152) | 0.398**<br>(0.150) |
| Occupancy Rate | 0.814***<br>(0.174) | 0.792***<br>(0.172) | 0.829***<br>(0.174) | 0.804***<br>(0.172) |
| 25-50 beds | 0<br>(.) | 0<br>(.) | 0<br>(.) | 0<br>(.) |
| 50-100 beds | 1.204<br>(0.669) | 1.171<br>(0.661) | 1.107<br>(0.670) | 1.104<br>(0.662) |
| 100-150 beds | 2.220***<br>(0.658) | 2.199***<br>(0.650) | 2.162**<br>(0.659) | 2.145**<br>(0.651) |
| 150-200 beds | 2.682***<br>(0.712) | 2.599***<br>(0.704) | 2.516***<br>(0.713) | 2.491***<br>(0.704) |
| 200+ beds | 2.077**<br>(0.805) | 1.997*<br>(0.795) | 1.948*<br>(0.807) | 1.924*<br>(0.797) |
| Constant | 1.995**<br>(0.684) | 1.846**<br>(0.676) | 2.116**<br>(0.684) | 1.923**<br>(0.677) |
| Fixed Effects | County | County | County | County |
| Depvar mean | 4.488 | 4.488 | 4.488 | 4.488 |
| Adj R2 | 0.34 | 0.35 | 0.33 | 0.35 |
| N | 2038 | 2038 | 2038 | 2038 |

Another interpretation is that these results are driven by the individual risk factors of staff members themselves, rather than their neighborhoods. This could have different policy implications, such as suggesting that nursing homes could potentially be protected if we provided staff with non-public transportation options. However, data from the American Community Survey suggests that very few (less than 5%) nursing home workers take public transportation to work in the study states (S2 Table). Furthermore, after controlling for the overall neighborhood measure, there is no effect of a measure of neighborhood public transportation use that is restricted to workers in the education and health care industry (S3 Table, column 1). Likewise, after controlling for the neighborhood racial composition, there is no effect of the share of workers who work on the same block as the nursing home who are non-white, which should be a better proxy for the racial composition of a nursing home’s staff (S3 Table, column 2).

It is possible that the initial infection of a facility and the containment or spread of the virus at the facility are affected by different factors. To study this, S4 Table reproduces the main results using a binary indicator for an outbreak, whether or not a facility reported any death, rather than the continuous measure used throughout the paper. Columns (2) and (3) show that the main results apply when investigating the presence of an outbreak: staff neighborhood characteristics continue to be one of the most important predictors of facility infection, and there continues to be a large effect of for-profit status. There are some differences: the effect of chain status is not significant here, and there is a slightly negative effect of star rating on the binary measure, and a positive effect of the resident non-white share, suggesting that lower-rated facilities and facilities with more non-white residents were more likely to experience an outbreak, even though outbreak size was not correlated with these characteristics.

These results beg the question: what types of facilities are likely to have staff who live in more dense and nonwhite neighborhoods with more public transportation? Are lower-quality, for-profit, nursing homes more likely to have higher staff neighborhood exposure? Are the most exposed facilities also the ones with the lowest wages? Figure 4 shows partial correlations of the staff neighborhood measures with other facility characteristics, controlling for county fixed effects, with a particular focus on measures related to a facility’s staffing practices. Both staff neighborhood public transportation use and staff neighborhood non-white share are positively correlated with larger, for-profit, and lower-rated facilities, but the correlations are relatively small. Likewise, there is a small negative correlation of the wage paid to certified nursing assistants (40% of the nursing home workforce, and the occupation likely to have the most contact with patients). On the other hand, staff exposure exhibits much larger correlations with the demographics of the residents: facilities with more Medicaid patients, and especially more nonwhite patients, are more likely to have higher measures of the staff neighborhood exposure measures. These results explain why in Table 1, the resident nonwhite share coefficient becomes insignificant after including the staff neighborhood measures, and suggest a potential channel for observed racial disparities in COVID infection across nursing homes: nursing homes with more non-white residents appear to employ more staff from the most highly exposed neighborhoods.

**Figure 4:**
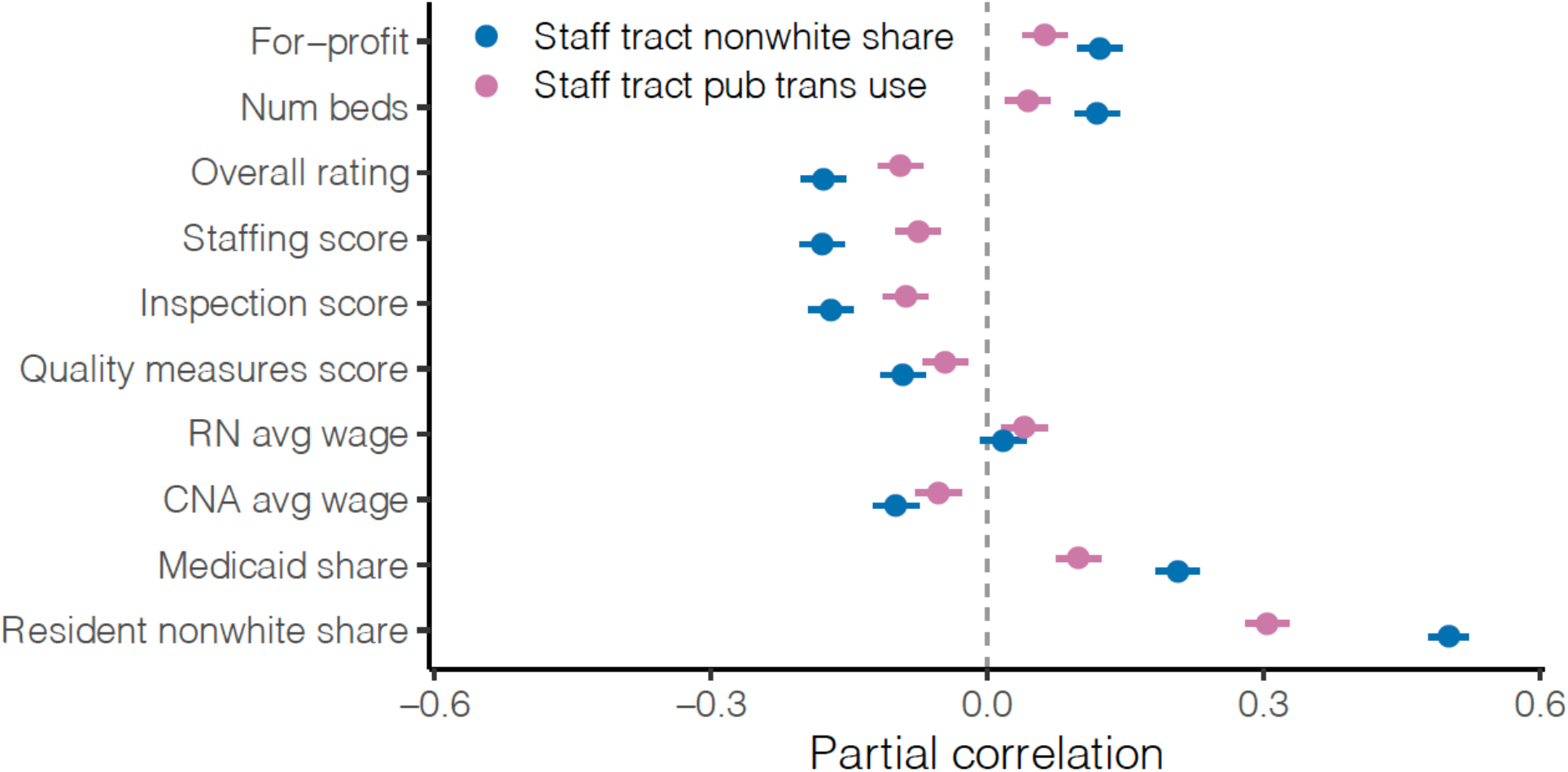
Correlates of staff neighborhood white share, controlling for county fixed effects. This figure reports partial correlations of different facility characteristics listed along the y-axis with two neighborhood characteristics of interest: public transportation use and non-white share. Correlations control for county fixed effects. Lines represent 95% confidence intervals.

## Discussion

This study finds that even after controlling for a nursing home’s county, nursing homes whose staff come from denser, less white neighborhoods with more public transportation use have had significantly larger outbreaks of COVID-19, and that these measures are much more powerful in explaining differences in death rates within a county than many other facility characteristics (such as nursing home rating), and also than the same characteristics of the nursing home’s own neighborhood.

While there were early efforts to close nursing homes to visitors and protect nursing home residents, the experience of these homes has indicated that these efforts were not nearly enough, with significant numbers of homes becoming infected after they were “locked down.” Because of the close personal contact they have with residents, staff members are a likely source of transmission, and these results lend support to this hypothesis, and underscore the importance of policies such as PPE and frequent testing for nursing home staff going forward. However, the small effect of facility management variables compared to the large effect of staff neighborhood characteristics suggests that it may ultimately be necessary to control outbreaks in the community in order to control facility outbreaks. It is possible that these relationships are specific to the first wave of deaths in the spring and early summer of 2020, and that as nursing homes gained experience with controlling outbreaks, other variables became more important in determining deaths from COVID-19. While this study does not investigate deaths in the later months of the pandemic, descriptive evidence from other researchers appears to indicate that community spread has remained important. [9]

Previous research has documented substantial segregation in long-term care, and how the location of high-quality facilities may exacerbate other inequalities. [10, 11, 12, 13, 14] In the case of COVID-19, even though this study does not find evidence of a significant effect of facility rating on facility outbreaks, it does uncover the concerning finding that the facilities that employ staff from neighborhoods that are more exposed to COVID-19 infection are also the facilities that serve more non-white residents. Early evidence suggests that black and Latino communities have been hit hardest by the pandemic. [15, 16, 17, 18, 19] The fact that the nursing home industry draws staff disproportionately from these communities in general may explain some of the enormous impact of the pandemic on nursing homes.

Finally, the persistent and relatively large effect of for-profit status on COVID-19 outbreaks as well as the consistently small or zero effect of a facility’s star rating (after accounting for geography and other facility characteristics) both merit further study, as it suggests that non-profit homes may have responded differently to the pandemic, but that the rating system was not able to predict these differences. In addition, the fact that wages are not highly correlated with high staff exposure also offers an opportunity for future study to understand why certain homes employ more heavily from more exposed communities; it may indicate that these nursing homes are most conveniently located for people living in these communities, or that facilities with more and less non-white residents have different hiring practices.

This study has a few limitations. First, it is important to note that the analysis in this study is correlational and there may therefore be omitted variables that are driving the results. For example, S5 Table shows that the coefficients on staff neighborhood characteristics are reduced if the distance of the nursing home to the central business district of the nearest metropolitan area is included, though they remain relatively large and statistically significant. This could either mean that (1) staff neighborhood characteristics are a true risk factor for facility outbreaks, and nursing homes that are more centrally located are simply likelier to draw staff from more exposed neighborhoods, or (2) centrality affects facility outbreaks through other mechanisms besides staff neighborhoods. However, in this case, the fact that staff neighborhood characteristics continue to be significant after controlling for centrality suggest that the former may be more important, and also offer a lower bound for these effects. A second limitation is that because the analysis is within counties, it cannot offer much insight into the effects of different county- or state-level policies in the COVID-19 response, though these are likely to have been important in determining infection rates and deaths. Third, although Table 2 offers suggestive evidence that the relationship between staff neighborhood characteristics and can be explained by higher infection rates in these neighborhoods, it is limited by the lack of data on infection rates by neighborhood (only one state in the sample provides case-level data at the tract-level, the remaining states in Table 2 provided data at the town or zip level, and the other states not included in Table 2 only report case data at the county level). More granular data of this form would help confirm the hypotheses of this study.

## Conclusions

During the first wave of the pandemic, which nursing homes experienced the largest outbreaks of COVID-19 within a county was not random, but it was also not largely determined by other measures of quality commonly cited in the nursing home literature, such as star rating. Instead, a key determinant was the characteristics of the neighborhoods where nursing home staff members lived—facilities whose staff lived in denser, less white neighborhoods with more public transportation use have had significantly more deaths than other facilities in the same county.

## Data Availability

Will be made public on Github

## Acknowledgements

I am very grateful for comments and discussions with David Cutler, Leemore Dafny, Ed Glaeser, Ashvin Gandhi, and Larry Katz that contributed to the ideas in this paper.

