## Appendix for "Relationship between nursing home COVID-19 outbreaks and staff neighborhood characteristics"

### Data appendix

**State COVID-19 data**

We obtain publicly released facility-level data on COVID-19 infections from 18 states. We began collecting this data in mid-April and continued to do so approximately every week until the week of July 10, 2020. Since some states do not report facilities with "closed outbreaks"'--i.e. facilities with no current cases--we use the historical data to build a cumulative measure of whether a facility was ever infected as if they appeared on any list. These data should largely reflect all nursing homes that have ever reported a COVID-19 infection, though data is usually self-reported by facilities and may contain errors, and states also differ in the exact data that they report. Notably, Maryland only reported facilities that had cases after April 15, New York only reports deaths that occurred at the nursing home (rather than all deaths among nursing home residents), and four states only report nursing homes with 2+ cases. We matched the names on these lists to the administrative data on the universe of nursing homes. This allows us to calculate the number of deaths per bed at each nursing home, and to compare characteristics of nursing homes with low and high numbers of deaths. In terms of numbers of deaths, the 18 states in our sample represent over 80% of the total deaths from COVID at the time of data collection and contain all of the top 10 states. The states with many deaths for whom we do not have facility-level data include Texas, Ohio, Indiana, Arizona, Texas, and Virginia.

Eight of our sample states also released easily accessible data on confirmed or probable COVID-19 cases at a finer geography than county. These states and the lowest level of geography at which they supply data on cases were: Connecticut (town), Florida (town), Illinois (zip), Louisiana (tract), Massachusetts (town), Maryland (zip), Rhode Island (zip), and South Carolina (zip). For all states except Louisiana, we use the case rates as of the date the nursing home infection data was pulled. For Louisiana, we use data from May 31, because we have not been able to obtain data from the previous week.

**Nursing home staff geography measures**

To calculate our nursing home staff neighborhood characteristics, we first use the 2017 LODES Workplace Area Characteristics data to identify the nursing home's census block. This data records the number of workers in the education and health care sector who work on a given census block. We geocode the nursing home addresses, and compute a predicted number of nursing home workers based on the national ratio of workers to residents (1.6 million workers to 1.3 million residents), and the average number of residents for each facility from the Nursing Home Compare data. For 83% of facilities, health care employment on the coded census block is more than half the predicted employment, and we consider this a match. For the remaining 11% of facilities, we use the closest census block in the block group that meets this condition. This allows us to match an additionall 11% of facilities, leaving about 511 facilities unmatched, which we exclude from our sample.

Using the census block chosen above, we then use the LODES Origin-Destination (OD) data to identify the home census blocks of workers who work on the same census block as the nursing home and belong to the "all other services'" industry group. One concern is that there may be many more service sector employers that are on the same census block as the nursing home, and these employers have staff who live in completely different neighborhoods than the nursing home's staff. We find on the median block, 92% (IQR: [.62, 1]) of the service employment on these census blocks is in the education and health care sector. This gives us reassurance that we are not mostly picking up an entirely different type of employer on these blocks and that our geocoding is likely to be fairly accurate.

However, it is still possible that there are other education or health-care employers on the same block as the nursing home. One particular case of this is nursing homes located near hospitals. Indeed, we do find some blocks with unreasonably large numbers of health care workers for a nursing home (5000+). However, in general, we find that the total employment numbers are reasonable. Using the calculation for predicted nursing home employment above, the median block in our sample has an actual service sector employment to predicted employment ratio of 1.5 (IQR [1.1, 2.7]). Thus, we believe it is likely that the measured neighborhood characteristics will be largely representative of the types of neighborhoods where a facility's employees are likely to live.

To calculate the "share frontline" measure, we define a frontline worker as a worker in an essential industry (as defined in Tomer and Kane, 2020) in an occupation that cannot work from home (as defined in Dingel and Neiman, 2020).

### Appendix 1: Data Appendix

**State COVID-19 data**
We obtain publicly released facility-level data on COVID-19 infections from 18 states. We began collecting this data in mid-April and continued to do so approximately every week until the week of July 10, 2020. Since some states do not report facilities with “closed outbreaks”'--i.e. facilities with no current cases--we use the historical data to build a cumulative measure of whether a facility was ever infected as if they appeared on any list. These data should largely reflect all nursing homes that have ever reported a COVID-19 infection, though data is usually self-reported by facilities and may contain errors, and states also differ in the exact data that they report. Notably, Maryland only reported facilities that had cases after April 15, four states only report nursing homes with 2+ cases. We matched the names on these lists to the administrative data on the universe of nursing homes. This allows us to calculate the share of nursing homes that have been infected, as well as compare characteristics of infected and non-infected homes. In terms of numbers of deaths, the 18 states in our sample represent over 80% of the total deaths from COVID so far and contain all of the top 10 states. The states with many deaths for whom we do not have facility-level data include Texas, Ohio, Indiana, Arizona, Texas, and Virginia.

Eight of our sample states also released easily accessible data on confirmed or probable COVID-19 cases at a finer geography than county. These states and the lowest level of geography at which they supply data on cases were: Connecticut (town), Florida (town), Illinois (zip), Louisiana (tract), Massachusetts (town), Maryland (zip), Rhode Island (zip), and South Carolina (zip). For all states except Louisiana, we use the case rates as of the date the nursing home infection data was pulled. For Louisiana, we use data from May 31, because we have not been able to obtain data from the previous week.

**Nursing home staff geography measures**
To calculate our nursing home staff neighborhood characteristics, we first use the 2017 LODES Workplace Area Characteristics data to identify the nursing home's census block. This data records the number of workers in the education and health care sector who work on a given census block. We geocode the nursing home addresses, and compute a predicted number of nursing home workers based on the national ratio of workers to residents (1.6 million workers to 1.3 million residents), and the average number of residents for each facility from the Nursing Home Compare data. For 83% of facilities, health care employment on the coded census block is more than half the predicted employment, and we consider this a match. For the remaining 11% of facilities, we use the closest census block in the block group that meets this condition. This allows us to match an additional1 11% of facilities, leaving about 511 facilities unmatched, which we exclude from our sample.

Using the census block chosen above, we then use the LODES Origin-Destination (OD) data to identify the home census blocks of workers who work on the same census block as the nursing home and belong to the “all other services"” industry group. One concern is that there may be many more service sector employers that are on the same census block as the nursing home, and these employers have staff who live in completely different neighborhoods than the nursing home's staff. We find on the median block, 92% (IQR: [.62, 1]) of the service employment on these census blocks is in the education and health care sector. This gives us reassurance that we are not mostly picking up an entirely different type of employer on these blocks and that our geocoding is likely to be fairly accurate.

However, it is still possible that there are other education or health-care employers on the same block as the nursing home. One particular case of this is nursing homes located near hospitals. Indeed, we do find some blocks with unreasonably large numbers of health care workers for a nursing home (5000+). However, in general, we find that the total employment numbers are reasonable. Using the calculation for predicted nursing home employment above, the median block in our sample has an actual service sector employment to predicted employment ratio of 1.5 (IQR [1.1, 2.7]). Thus, we believe it is likely that the measured neighborhood characteristics will be largely representative of the types of neighborhoods where a facility's employees are likely to live.

To calculate the “share frontline” measure, we define a frontline worker as a worker in an essential industry (as defined in Tomer and Kane, 2020) in an occupation that cannot work from home (as defined in Dingel and Neiman, 2020).

**S1 Table: Summary statistics for analysis sample**

| Variable | Mean | Std. Dev. |
|---|---|---|
| Number of beds | 119 | 67 |
| For-profit | .73 | |
| Non-profit | .24 | |
| Public | .03 | |
| Chain | .58 | |
| Star rating | 3.1 | 1.4 |
| - Inspection rating | 2.7 | 1.2 |
| - Staffing rating | 2.9 | 1.1 |
| - Quality measure rating | 3.8 | 1.2 |
| RN wage | 34.3 | 6.4 |
| CNA wage | 15.0 | 2.7 |
| Occupancy rate | .84 | .13 |
| Medicaid share | .60 | .23 |
| Resident share non-white | .23 | .23 |
| Staff tract pop density (pp/sq mi) | 4906 | 7386 |
| Staff tract pub trans use | .05 | .08 |
| Staff tract share nonwhite | .28 | .17 |
| Staff tract pov rate | .19 | .07 |
| Staff tract share frontline | .31 | .03 |
| NH tract pop density (pp/sq mi) | 4543 | 9484 |
| NH tract pub trans use | .05 | .10 |
| NH tract share nonwhite | .25 | .22 |
| NH tract pov rate | .18 | .17 |
| NH tract share frontline | .30 | .06 |

**S2 Table: Characteristics of nursing home workers from American Community Survey and BLS data**

| Top Occupations (OES) | Share |
|---|---|
| - Certified Nursing Assistant | .38 |
| - Licensed Practical Nurses | .13 |
| - Registered Nurses | .10 |
| - Food preparation and serving | .10 |
| - Building cleaners | .10 |
| - Office and administrative support | .05 |
| - Other healthcare practitioners (therapists, etc.) | .03 |
| - Laundry workers | .02 |
| - Other | .09 |
| Demographics (ACS) | |
| - Female | .84 |
| - White non-hispanic | .55 |
| - Black | .27 |
| - Hispanic or Latino | .06 |
| - High school or less | .38 |
| - Some college | .27 |
| - Two year degree | .15 |
| - Four year degree or more | .19 |
| - Commute to work by car | .92 |
| - Commute to work on public transportation | .04 |
| - Annual wage/salary income ≤ \$30,000 | .59 |
| - Annual wage/salary income ≤ \$50,000 | .82 |

**S3 Table: Relationship of facility infection with staff characteristics compared to staff neighborhood characteristics**

| | (1) | (2) |
|---|---|---|
| Staff tract pubtrans use | 1.159*** | |
| | (0.266) | |
| Staff tract PT share (Ed/Health) | 0.0465 | |
| | (0.139) | |
| Staff tract share nonwhite | | 1.078*** |
| | | (0.317) |
| Staff on block share nonwhite | | -0.238 |
| | | (0.208) |
| For-profit | 0.518** | 0.541** |
| | (0.193) | (0.193) |
| Chain | 0.375* | 0.363* |
| | (0.163) | (0.163) |
| Star rating | 0.0476 | 0.0470 |
| | (0.0915) | (0.0916) |
| No prior infection viol. | 0.254 | 0.253 |
| | (0.195) | (0.196) |
| Medicaid share | -0.00697 | -0.00784 |
| | (0.0880) | (0.0880) |
| Resident share nonwhite | -0.214 | -0.295* |
| | (0.121) | (0.130) |
| Avg severity | -0.0597 | -0.0516 |
| | (0.0813) | (0.0817) |
| Occupancy Rate | 0.664*** | 0.672*** |
| | (0.0871) | (0.0871) |
| 25-50 beds | 0 | 0 |
| | (.) | (.) |
| 50-100 beds | 0.540 | 0.531 |
| | (0.328) | (0.328) |
| 100-150 beds | 1.109*** | 1.115*** |
| | (0.333) | (0.333) |
| 150-200 beds | 1.732*** | 1.717*** |
| | (0.368) | (0.368) |
| 200+ beds | 1.163** | 1.155** |
| | (0.409) | (0.409) |
| Constant | 2.053*** | 2.037*** |
| | (0.343) | (0.344) |
| fe | County | County |
| ymean | 3.735 | 3.735 |
| r2_a | 0.29 | 0.29 |
| N | 6146 | 6146 |

Standard errors in parentheses
$^{*}$ $p < .05$, $^{**}$ $p < .01$, $^{***}$ $p < .001$

**S4 Table: Relationship of binary measure of facility infection (any death) with facility and neighborhood characteristics**

| | (1) | (2) | (3) |
|---|---|---|---|
| Staff tract pubtrans use | | 0.0888*** | |
| | | (0.0249) | |
| NH tract pubtrans use | | -0.0245 | |
| | | (0.0127) | |
| Staff tract share nonwhite | | | 0.0638*** |
| | | | (0.0169) |
| NH tract share nonwhite | | | -0.0121 |
| | | | (0.00913) |
| For-profit | 0.0404** | 0.0396** | 0.0393** |
| | (0.0139) | (0.0139) | (0.0139) |
| Chain | 0.0183 | 0.0190 | 0.0189 |
| | (0.0117) | (0.0117) | (0.0117) |
| Overall Rating | -0.0131* | -0.0129* | -0.0124 |
| | (0.00658) | (0.00658) | (0.00659) |
| No prior infection viol. | 0.0119 | 0.0116 | 0.0114 |
| | (0.0141) | (0.0141) | (0.0141) |
| Medicaid share | -0.00554 | -0.00496 | -0.00517 |
| | (0.00653) | (0.00653) | (0.00652) |
| Resident share nonwhite | 0.0292*** | 0.0237** | 0.0173 |
| | (0.00845) | (0.00890) | (0.00992) |
| Avg severity | -0.0229*** | -0.0217*** | -0.0206*** |
| | (0.00612) | (0.00613) | (0.00615) |
| Occupancy Rate | 0.0208** | 0.0203** | 0.0208** |
| | (0.00636) | (0.00636) | (0.00635) |
| 25-50 beds | 0 | 0 | 0 |
| | (.) | (.) | (.) |
| 50-100 beds | 0.127*** | 0.130*** | 0.129*** |
| | (0.0236) | (0.0236) | (0.0236) |
| 100-150 beds | 0.231*** | 0.233*** | 0.234*** |
| | (0.0240) | (0.0240) | (0.0240) |
| 150-200 beds | 0.301*** | 0.301*** | 0.300*** |
| | (0.0265) | (0.0265) | (0.0265) |
| 200+ beds | 0.326*** | 0.325*** | 0.324*** |
| | (0.0295) | (0.0294) | (0.0294) |
| Constant | 0.208*** | 0.207*** | 0.209*** |
| | (0.0247) | (0.0247) | (0.0247) |
| Fixed Effects | County | County | County |
| Depvar mean | 0.455 | 0.455 | 0.455 |
| Adj R2 | 0.40 | 0.40 | 0.40 |
| N | 6132 | 6132 | 6132 |

Standard errors in parentheses

* $p < .05$, ** $p < .01$, *** $p < .001$

**S5 Table: Relationship of facility deaths per bed with distance to central business district and staff and nursing home neighborhood characteristics**

| | (1) | (2) |
|---|---|---|
| Distance to CBD | -0.978*** | -1.052*** |
| | (0.251) | (0.244) |
| Staff tract pubtrans use | 0.799* | |
| | (0.336) | |
| NH tract pubtrans use | -0.0297 | |
| | (0.163) | |
| Staff tract share nonwhite | | 0.569* |
| | | (0.244) |
| NH tract share nonwhite | | -0.0255 |
| | | (0.126) |
| For-profit | 0.529** | 0.530** |
| | (0.193) | (0.193) |
| Chain | 0.374* | 0.370* |
| | (0.162) | (0.162) |
| Star rating | 0.0500 | 0.0504 |
| | (0.0915) | (0.0916) |
| No prior infection viol. | 0.263 | 0.264 |
| | (0.195) | (0.195) |
| Medicaid share | 0.0357 | 0.0291 |
| | (0.0885) | (0.0884) |
| Resident share nonwhite | -0.290* | -0.345* |
| | (0.123) | (0.137) |
| Avg severity | -0.0673 | -0.0613 |
| | (0.0813) | (0.0816) |
| Occupancy Rate | 0.656*** | 0.660*** |
| | (0.0876) | (0.0876) |
| 25-50 beds | 0 | 0 |
| | (.) | (.) |
| 50-100 beds | 0.537 | 0.528 |
| | (0.328) | (0.328) |
| 100-150 beds | 1.116*** | 1.114*** |
| | (0.333) | (0.333) |
| 150-200 beds | 1.680*** | 1.663*** |
| | (0.369) | (0.368) |
| 200+ beds | 1.117** | 1.096** |
| | (0.409) | (0.409) |
| Constant | 2.092*** | 2.095*** |
| | (0.343) | (0.343) |
| fe | County | County |
| ymean | 3.736 | 3.737 |
| r2_a | 0.29 | 0.29 |
| N | 6141 | 6142 |
